# Finding Case Report Nuggets: A Web-Based Tool for Mining and Enhancing the Value of Clinical Case Reports

**DOI:** 10.1101/2025.11.13.25340162

**Authors:** Arthur W. Holt, Neil R. Smalheiser

## Abstract

**Objectives:** Case reports are eyewitness reports of medical phenomena, such as adverse effects of treatments, outcomes of new surgical techniques, descriptions of rare diseases, unusual presentations of common diseases, or emerging infectious outbreaks. Although any single case report may be confounded, biased or erroneous, observations that are separately reported in multiple independent publications are more likely to be reliable, and so the accumulated evidence should have more value than any single report on its own. This notion led us to analyze the case reports literature in search of nuggets: collections of multiple case reports that describe similar main findings.

**Materials and Methods:** To identify nuggets in collections of case reports retrieved in PubMed queries, semantic similarities among the case reports were computed based on titles and main finding sentences extracted from the abstracts, and then grouped into communities with a graph database. The initial communities were then merged with a secondary hierarchical clustering process.

**Results:** Computed nuggets of size 4-100 articles are displayed along with large language model (LLM)-computed summaries, the title of the nugget’s central article, and hyperlinks for viewing as well as export to our companion tool Anne O’Tate for further analysis. A variety of advanced options are also offered; users can optionally submit feedback on the quality of computed nuggets.

**Discussion:** Our free, public tool https://arrowsmith.psych.uic.edu/casereports facilitates the identification of nuggets and their summarization and mining. This should enhance the value of case report evidence and assist clinicians as well as those performing evidence syntheses of the published literature.

## BACKGROUND AND SIGNIFICANCE

Clinical case reports comprise a significant portion of the peer-reviewed biomedical literature: Over 2.4 million articles are indexed as Case Reports in PubMed, or roughly 15% of the total publications. Case reports are eyewitness reports of medical phenomena, often reporting on timely subjects such as adverse effects of treatments, outcomes of new surgical techniques, descriptions of rare diseases (or unusual presentations of common diseases), or emerging infectious outbreaks. Because they are uncontrolled observations, case reports are relegated to the bottom of the Evidence Hierarchy and often excluded in evidence syntheses that focus preferentially on randomized controlled trials and controlled observational studies such as case-control and cohort studies.

Nevertheless, the medical value of case reports is undeniable.[1–9] For example, most clinical trials describe only the short-term effects of an intervention, whereas longer-term adverse effects or abuse of medications are likely to be described in case reports.[10–12] The first publications to report the existence of AIDS were three case reports. Moreover, case reports are useful adjuncts in medical education.[13, 14]

Although any single case report may be confounded, biased or erroneous,[15] observations that are separately reported in multiple independent publications are more likely to be reliable,[16] and so the accumulated evidence should have more medical value than any single report on its own. A number of approaches have aggregated evidence across groups of case reports, including semantic annotation of rare disease phenotypes [17] and consensus clustering of adverse drug events.[18]

Our own group analyzed the case reports literature in search of “nuggets”, collections of case reports that describe similar observations, outcomes, or main findings.[19–21] We found that roughly 1/7th of topics searched in PubMed retrieved a set of case reports containing at least one nugget made up of four or more articles.[19] In prior work, we found that the title of a clinical case report explicitly states the main finding in the overwhelming majority of articles [19–21]. We also created manually annotated corpora of case reports and found that most articles restate the main finding in a sentence (usually the first or last) within the article abstract [19–21]. Machine-learning methods were devised to extract the main finding sentence(s) from the article abstract.[20, 21]

The field of biomedical information retrieval has extensively explored how to cluster articles according to topical similarity and relevance, with or without employing topic modeling;[22–34] Although some previous web tools have implemented displays of similar articles, to our knowledge, only BioTextQuest v2.0 is currently maintained.[35] In the present paper, we consider how to find nuggets in an automated manner within any set of articles retrieved from PubMed. It should be emphasized that finding nuggets involves clustering articles that share similar main findings, rather than sharing topical similarity in general. We describe the construction of a free, public, web-based tool, **Finding Case Report Nuggets**, that facilitates their identification, summarization and mining, and offer several use cases for clinicians, systematic reviewers and other biomedical end-users.

## MATERIALS AND METHODS

### Definitions

#### Nugget

A collection of case reports that describe similar or closely related observations, outcomes, or main findings.

#### Community

A group or cluster of nodes in a graph database as determined by the Louvain method of community detection.[36] In this paper, communities are detected and merged in order to identify nuggets.

#### Vector

A 768 dimensional embedding vector for titles and main findings as sentences, computed with the pre-trained PubMedBERT model.[37]

#### Similarity

The cosine of the angle between two title or main finding sentence vectors. Values range from −1 to 1, with 1 being interpreted as identical meaning.

#### Graph Database

A specialized database format that stores nodes, edges, and properties, that facilities the analysis of relationships between entities. For case reports, each article in the query result set is a node in the graph database. Title and main finding similarities define the connections (edges) between the nodes as well as a weight property used for community detection.

### Case report article corpus

Case report articles were retrieved from PubMed.gov in November 2024 with a query that selects 2,484,572 articles indexed by the NLM as Case Reports publication type. As well, to increase recall, we also included publications indexed as Letter that mention the word case or report in the metadata: (“case reports“[Publication Type]) OR (“letter“[Publication Type] AND (“Case“[All] or “Report“[All])). Also, articles with “case series” in the title added an additional 23,714 articles. Additionally, 222,658 articles, predicted to be case reports by the MultiTagger 1.0 model,[38] were added for a total of 2,730,944 articles to consider for nuggets. This list is updated weekly with new case reports from these sources.

### Case report feature derivation

For each case report article the attributes shown in Table 1 were retrieved from PubMed, processed with external embedding models, and stored in a lookup table to enable rapid pairwise comparisons among articles within a topical query set. As shown below, by far the most important feature was the title similarity. However, since not every article expresses its main finding in its title, we sought to increase recall by a logistic regression-based Main Finding model identifying the most likely main finding sentence from the article abstract.[21], and computing pairwise both title-main finding sentence similarity and main finding sentence-main finding sentence similarity across articles. Vector representations of 768 dimensions for both the article title and main finding sentence were computed with a BERT-based model that has been fine-tuned with PubMed literature.[37]

**Table 1.** Case Report Article Attributes.

| Article Feature | Source/Derivation |
| --- | --- |
| Title | Article Title as provided by PubMed XML metadata |
| Abstract | Abstract as provided by PubMed XML metadata |
| Main Finding | Main finding sentence extracted from the article abstract as determined by a rule-based model.[19] |
| Title PubMedBERT Embedding Vector | Embedding vector for the article title.[34] |
| Main Finding PubMedBERT Embedding Vector | Embedding vector for the article main finding sentence.[34] |

### Nugget detection overview

In general terms, our process for finding nuggets includes finding articles with similar main findings, and grouping them in two stages. For computational efficiency, a list of all articles from our case report corpus was retrieved and stored in a relational database with approximately 2.7 million articles. Semantic vectors for the titles, abstracts, and main finding sentences were pre-computed and stored in the database as well. During real-time usage of the tool, the user’s PubMed query result is matched to the pre-computed table of case reports and the related numeric vectors retrieved. As shown in Figure 1, similarities among the case reports are computed and then grouped into communities with a graph database. The initial communities are then merged with a secondary hierarchical clustering process. The resulting nuggets are then filtered by the user’s nugget size constraints and output to the web tool.

**Figure 1.**
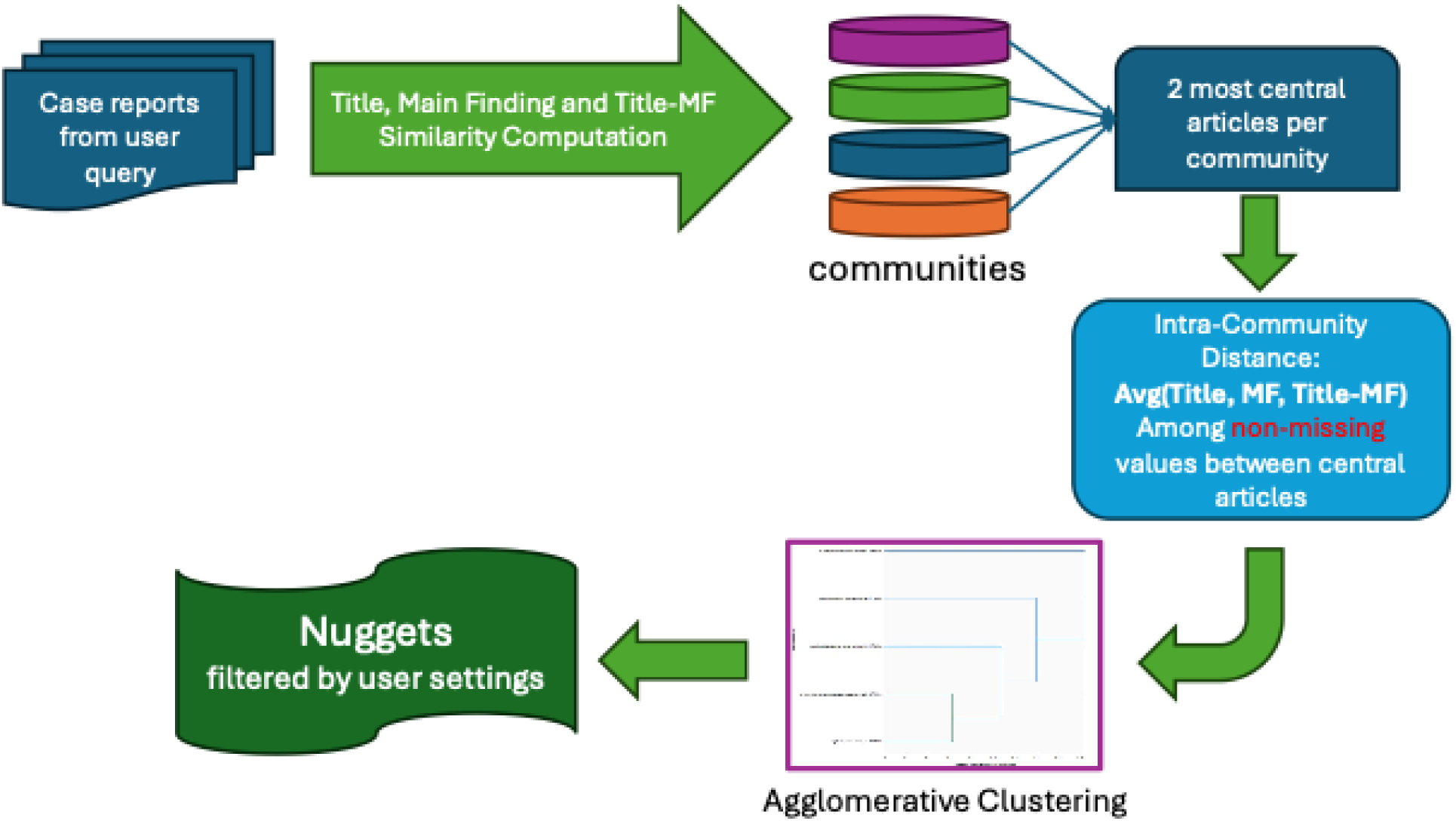
Nugget Detection Process Summary. Overview of the nugget determination process. The user query is executed with PubMed.gov, restricted to case reports, and processed with a two-stage merging algorithm to find nuggets with similar topics or main findings.

### Positive and negative PubMed queries used for tuning the model

In order to test experimental designs and optimize parameters, we examined 30 randomly chosen PubMed topical search queries, of which 17 were observed to contain one or more nuggets (Table 2).[19] These were contrasted with 30 geographical query examples that were not expected to produce a significant number of nuggets, by randomly choosing 30 North American cities for PubMed queries consisting of the city name mentioned in the Affiliation field. Articles written by authors in a single city will cover a wide variety of topics, but are not expected to be associated with a large number of similar case reports (apart from rare situations such as the early reports of AIDS which emerged from Haiti and New York City).

**Table 2.** Test Queries for Evaluating Nugget-Finding Algorithms.

| Topical Search Phrase | # Case Reports | # Nuggets |
| --- | --- | --- |
| Abbreviations as topic[MESH] | 28 | 0 |
| Annexin A5[MESH] | 25 | 0 |
| Bone Morphogenetic Protein 4[MESH] | 30 | 0 |
| Commotio Cordis[MESH] | 38 | 2 |
| Glyoxylates[MESH] | 27 | 0 |
| Overlearning[MESH] | 16 | 0 |
| Oviposition[MESH] | 37 | 0 |
| Podophyllum[MESH] | 20 | 0 |
| Sexual Abstinence[MESH] | 46 | 1 |
| Widowhood[MESH] | 36 | 0 |
| Corneal Endothelial Cell Loss[MESH] | 58 | 3 |
| Flavin-Adenine Dinucleotide[MESH] | 28 | 0 |
| Growth Differentiation Factor 5[MESH] | 33 | 2 |
| Guanidine[MESH] AND Sexually Transmitted Diseases, Viral[MESH] | 38 | 1 |
| Patellofemoral Pain Syndrome[MESH] | 41 | 1 |
| Pyonephrosis[MESH] | 75 | 2 |
| Respiratory Care Units[MESH] | 16 | 0 |
| Arcuate Nucleus[MESH] | 29 | 0 |
| Bunolol[MESH] | 30 | 1 |
| Histone-Lysine N-Methyltransferase[MESH] | 623 | 9 |
| Hexosaminidase B[MESH] | 49 | 3 |
| Receptors, Natural Killer Cell[MESH] | 57 | 1 |
| Interleukin-1 Receptor-Associated Kinases[MESH] | 54 | 2 |
| src-Family Kinases[MESH] | 44 | 0 |
| Total Disc Replacement[MESH] | 116 | 1 |
| Plakophilins[MESH] | 59 | 2 |
| Clathrin[MESH] | 24 | 1 |
| Delayed Graft Function[MESH] | 88 | 2 |
| Cholic Acid[MESH] | 25 | 1 |
| HLA-B52 Antigen[MESH] | 17 | 0 |

Thirty topical queries were analyzed with our algorithm nugget detection methodology. Shown is the number of case reports retrieved by each query, and the number of detected nuggets, i.e., merged communities that contained four or more articles.

### Graph database

In preliminary studies, textual (word and n-gram) similarity measures were investigated to identify nuggets, but performance was disappointing and a graph database approach was found to be more effective. For each case report article contained in the results retrieved from a given PubMed.gov query, a node was created within an empty graph database. Edges between the article nodes were drawn when one of three distinct measures of similarity were greater than a specific threshold. The first involves computing the cosine similarity between PubMedBERT embeddings of both titles. The second calculates the cosine similarity between the main-finding sentences of the two articles, again using PubMedBERT embeddings. The third examines cross-article similarity by taking the maximum of the cosine similarities between one article’s title and the other’s main finding (and vice versa).

### Finding communities

From the graph database, communities of articles are designated using the Louvain method of community detection,[36] with a granularity setting of 1.2. In general terms, the Louvain method iteratively optimizes modularity, i.e. the quality of network divisions, starting with each article in its own community. This method of community detection is not deterministic since nodes, in this case articles, are selected in random order for optimizing modularity. In order to maintain reproducibility, we set the random seed for this process to a fixed value so that identical article sets produce exactly the same nugget results. There is a known limitation of the Louvain method in that small well-defined communities may not be detected in large dense networks; however, this did not appear to be a substantial problem with our granularity setting of 1.2 and given that our approach generally avoids creating extremely dense networks. For each community designated by the Louvain, the two most central articles, as determined by degree of centrality, were retained for merging communities.

Edges between article nodes are created when the cosine similarity between article titles is > 0.72, the main finding sentence similarity is > 0.82, or the maximum of cosine similarity between one article’s title with the other’s main finding sentence is > 0.77. The initial thresholds for drawing edges were determined by examining the maximum values generated across three of the geographically based negative queries (Kansas City, Houston, Tampa) with the assumption that these queries would create few, if any meaningful nuggets. Since the negative cases did not provide an obvious threshold for title similarity due to wide variability, the average of the title similarity among manually identified nuggets within Shiitake Mushroom and Zolpidem test queries (0.745) was used as a starting level. As shown in Table 3, we examined a range of possible thresholds from 0.64 to 0.8, and tabulated the proportion of positive vs. negative queries that generate more than one community. Based on this, we chose a community detection threshold of 0.72 as having the best trade-off between sensitivity and selectivity. For example, Figure 2 shows the resulting graph network for the articles returned for the Shiitake mushroom query.

**Figure 2.**
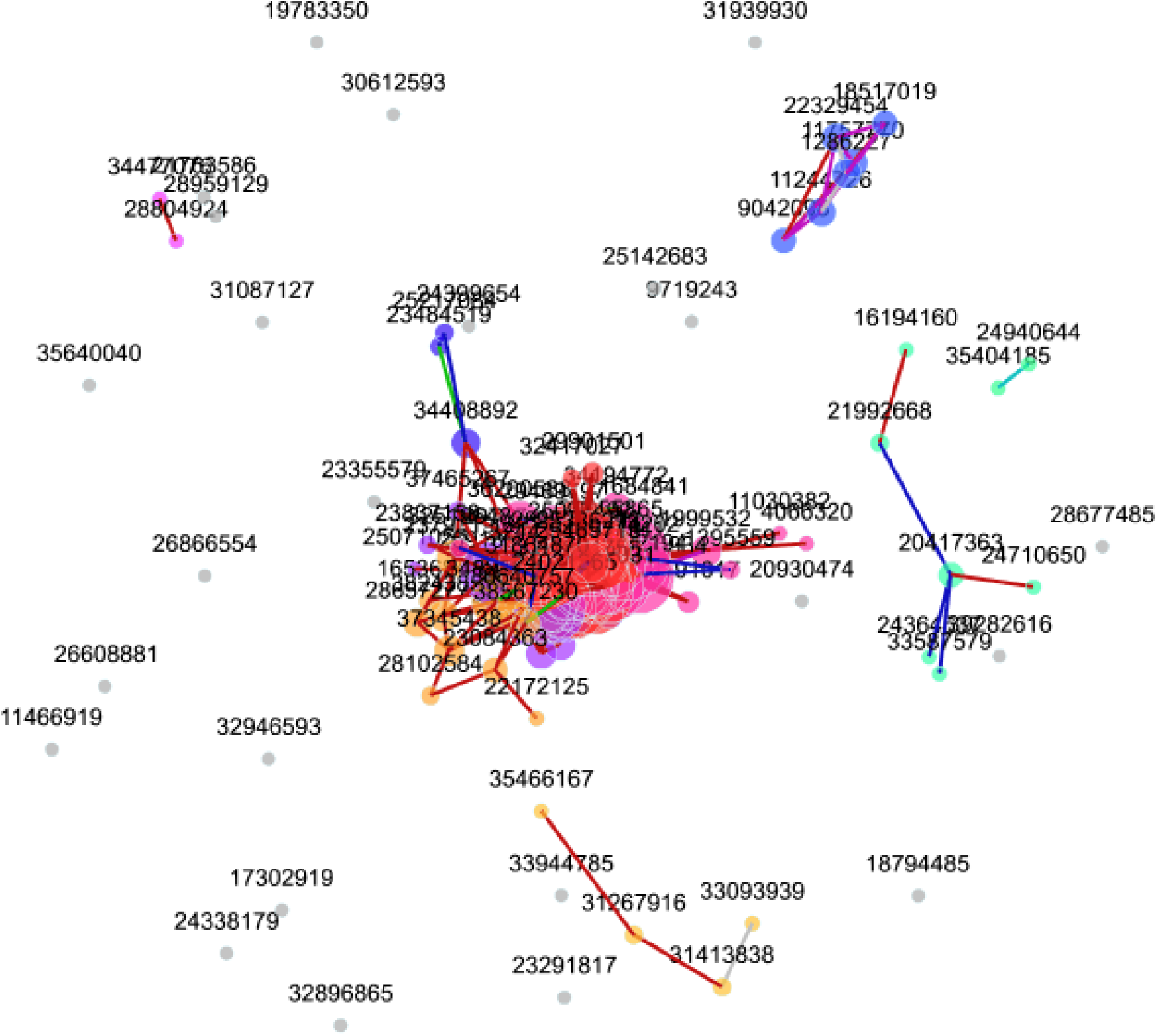
Example Graph for “Shiitake” PubMed Query. This network contains the 112 articles returned from a “Shiitake” PubMed query discussed in Results. Each article is a node and edges are drawn when one or more of three similarity metrics is greater than a fixed threshold (see Methods). Communities of articles are derived with the Louvain method.[33] Communities are shown with matching circle colors and significant similarities are shown as lines with red indicating title similarity, green indicating main finding similarity, and blue indicating title-to-main finding similarity. The large community in the center is comprised of articles describing shiitake-induced dermatitis cases arising from the ingestion of undercooked mushrooms. The isolated communities with 4 or more articles are related to other conditions such as bowel obstruction and pneumonitis (see Results).

**Table 3.**
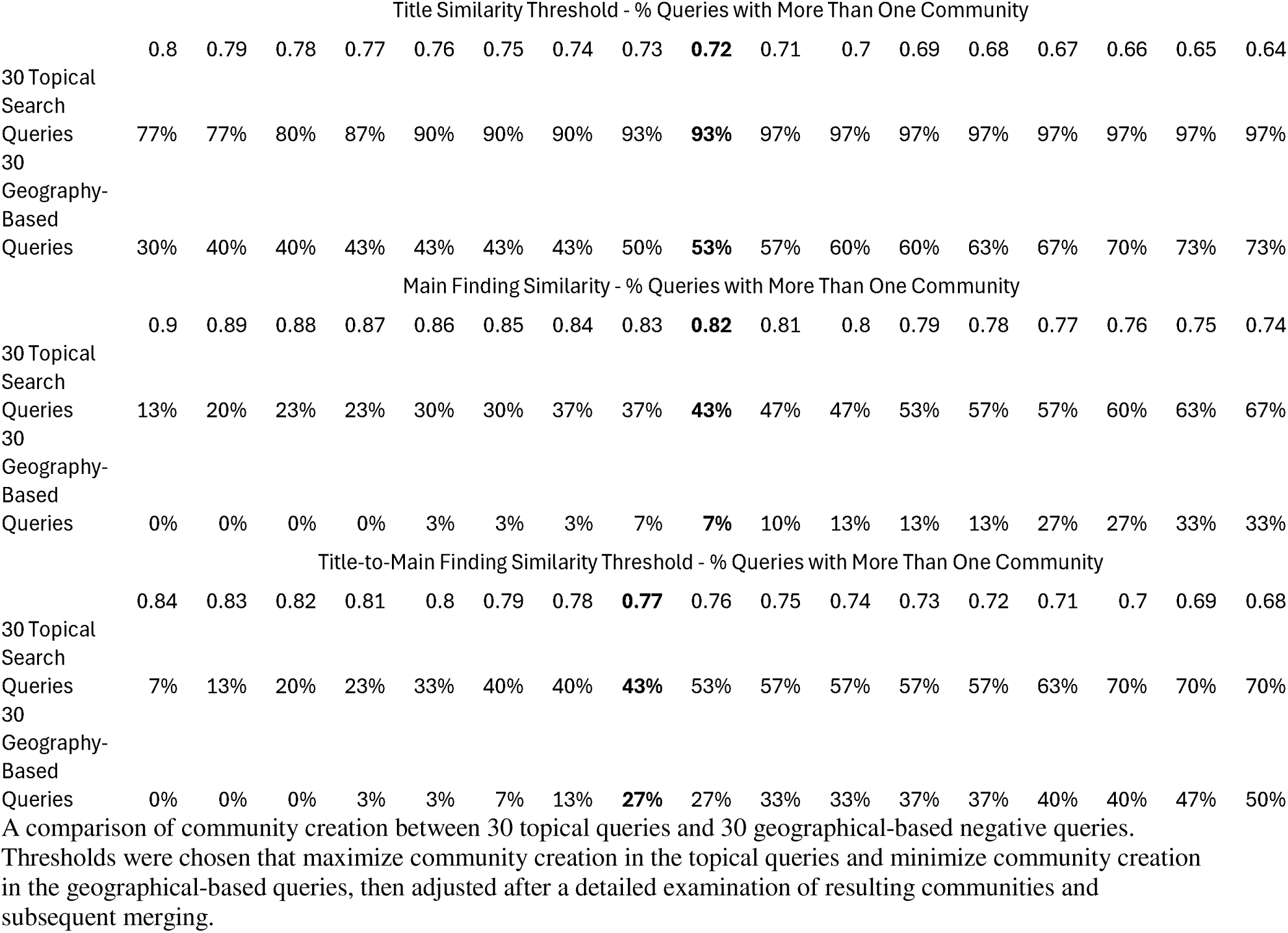
Sensitivity of Similarity Thresholds for Community Detection.

By far the most important feature for assigning similar pairs of articles was the criterion that cosine similarity between article titles is > 0.72. About 96.3% of all pairs within a nugget were identified based on similar titles. In contrast, only 2.3% of additional pairs were identified because their main finding sentence similarity is > 0.82, and only 1.4% of additional pairs because similarity between one article’s title with the other’s main finding sentence is > 0.77. Thus, although employing the main finding prediction model was helpful in augmenting the size of nuggets, it played only a minor role in the overall nugget finding process.

### Merging communities into nuggets

We implemented a secondary merging strategy in order to combine communities that discuss

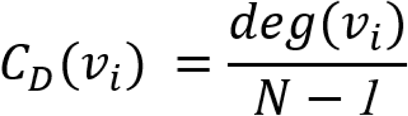

similar findings or topics in aggregate, but that were not connected directly by the Louvain method. For each initial community determined from the graph database, the two member articles v_1_ and v_2_ with the highest degrees of centrality are determined by undirected connections, given by:

where

C_D_ (v_i_) = degree centrality of node i
deg (v_i_) = number of edges connected to node i
N = total number of nodes in the graph

For each pairwise combination of the initial communities, a distance metric is calculated as follows for each of the 4 possible combinations of central articles between the 2 communities :

1. 1 - cosine similarity between embedding vectors for article titles between communities
2. 1 - cosine similarity between embedding vector for article title with embedding vector for the main finding of an article from the second community
3. 1 - cosine similarity between main finding embedding vectors between communities

This results in a maximum set of 12 distance metrics (i.e. 4 central article combinations x 3 similarity vector comparisons). A composite distance between each pair of communities is computed as the arithmetic mean of the 12 distances, excluding missing values generated as the result of missing main finding sentences for articles with no abstract available. Given the cosine function ranges from −1 to 1, our resulting distance metric ranges from 0 to 2, with larger values corresponding to less similarity. With these composite distance values, communities are merged with agglomerative clustering, complete linkage, with a merge cut-off of 0.42, corresponding to an average cosine similarity among central articles of 1 - 0.42, or 0.58. This threshold for merging was chosen by simulating levels ranging from an average cosine similarity of 0.3 to 0.7 across the topical queries, with particular focus on the Shiitake Mushroom and Zolpidem queries and a general motive to keep the cut-off as low as possible to avoid joining disparate communities. These simulations suggested that an optimal range was between .38 and .46, with .42 chosen after close examination of the resulting nuggets.

### Determining the minimum and maximum nugget size for display in the tool

We determined the minimum size of a computed merged community that should be deemed as a “nugget” for display by our tool. This was done quantitatively, by evaluating the difference between the distribution of merged community sizes in the 30 randomly chosen topical query set (i.e., a mixed set some of which contain nuggets) vs. the set of 30 geographically determined articles expected not to produce a significant number of nuggets. As shown in Figure 3, whereas 3.6% of the topical queries generated merged communities of 4 or more articles, only 0.2% of the negative queries generated merged communities of 4 or more articles. This is a striking difference, all the more striking given that the average number of PubMed articles retrieved by the negative queries (231) was several times greater than the average retrieved by the topical query set (60). This suggests that setting a threshold of 4 articles for a nugget would remove nearly all spurious clustering.

**Figure 3.**
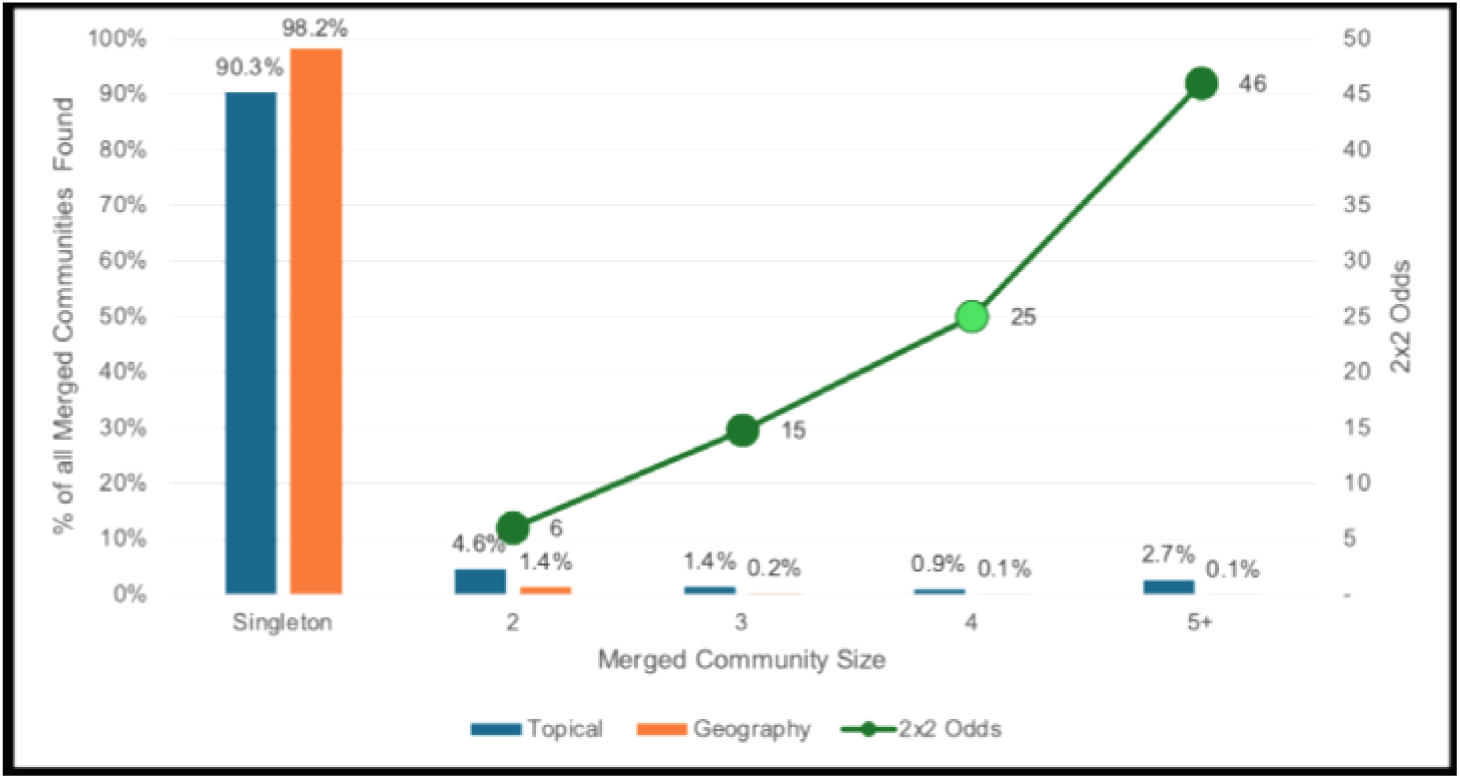
Comparison of Merged Community Sizes in Topical vs. Geographical Queries. The bars show the percentage of queries in the topical vs. geographical sets that gave rise to merged communities containing 2, 3, 4, or 5 or more articles. A comparison of the topical and geography community sizes, per a 2×2 contingency table contrasting the probability that merged communities are greater than or equal to the indicated size (green line), shows that 4 is the point of maximum inflection (shown in light green). This proves additional support for choosing a threshold of 4 articles for the default minimum size of a nugget.

When we tested topical PubMed queries retrieving large sets of articles (thousands rather than hundreds), sometimes very large nuggets were computed (>100 articles). Such nuggets had a tendency to be comprised of reports of increasing heterogeneity, discussing many different aspects within a similar topic or clinical problem rather than sharing specific main findings. We decided to cut off the display at 100 as a default setting for the tool (though the View Advanced Options button allows users to view nuggets of any size).

## RESULTS

### Online tool functionality

A publicly accessible web tool is provided at: https://arrowsmith.psych.uic.edu/casereports An editable PubMed Query box is provided in which an arbitrary search phrase may be entered. Upon clicking the Run Query button, the search phrase is passed to PubMed’s esearch API and the translated query is presented in the Query Translation box where it may be manually edited. The number of articles returned by PubMed is reported in the Query Information section. Several advanced search options may be unhidden with the “Show Advanced Options” button. The Show All Nuggets option releases the minimum and maximum nugget size criteria of 4 to 100. For convenience, an option to add publication date restriction is provided as well as an option to display the publication dates of the articles within a nugget, or to display the community merge structure resulting from the secondary merge process previously described.

### Nugget output

When the user clicks on the “Find Nuggets” button, a list of computed nuggets is displayed. For each nugget, the titles of the member articles have been summarized with a large language model (LLM), currently GPT-3.5,[39] with the system prompt of “You are summarizing groups of biomedical article titles in one very concise sentence” along with the user prompt “Summarize these article titles:” concatenated with each title on a separate line. The summary is presented in an LLM Summary column. Also displayed are the number of articles in each nugget, the title of the most central article as determined by the degree of centrality previously described, and a hyperlink to our Anne O’Tate tool for further mining and analysis.[40]

### Use cases and examples

A publicly accessible web tool is provided at: https://arrowsmith.psych.uic.edu/casereports An editable PubMed Query box is provided. Although the user can enter any arbitrary search phrase, we envision that a user will be searching for case reports related to a specific topic or phenomenon (e.g. bicycle injuries or mercury poisoning from seafood) rather than very general conditions associated with a very large literature (e.g. brain cancer). Here we present three examples of queries that an end-user might pose through the nugget tool:

The first one envisions a clinician, faced with a patient whose history is remarkable for ingestion of shiitake mushrooms, seeking to identify case report findings that are especially well-documented.[19] For the PubMed query “shiitake“[All Fields], there are 1,231 results, of which 112 articles are Case Reports. Reading or even skimming 112 articles would be cumbersome and would require great mental effort to discern recurring trends. However, entering this query into our tool (Figure 4) returns a simple and very informative output of 4 different nuggets (Figure 5). Of these, 65 articles present different aspects of a unique flagellate dermatitis associated with eating shiitake mushrooms, and a smaller nugget consists of 6 reports of this dermatitis in novel geographical settings. Four articles report on cases of small bowel obstruction, and six articles report on cases of hypersensitivity pneumonitis (Figure 6). Each of these nuggets is hyperlinked, which exports the list of articles to our sister tool Anne O’Tate,[40] which permits them to be mined further. Examining the set of articles in a nugget (e.g., Figs. 6, 7) confirms that they are, indeed, discussing similar main findings.

**Figure 4.**
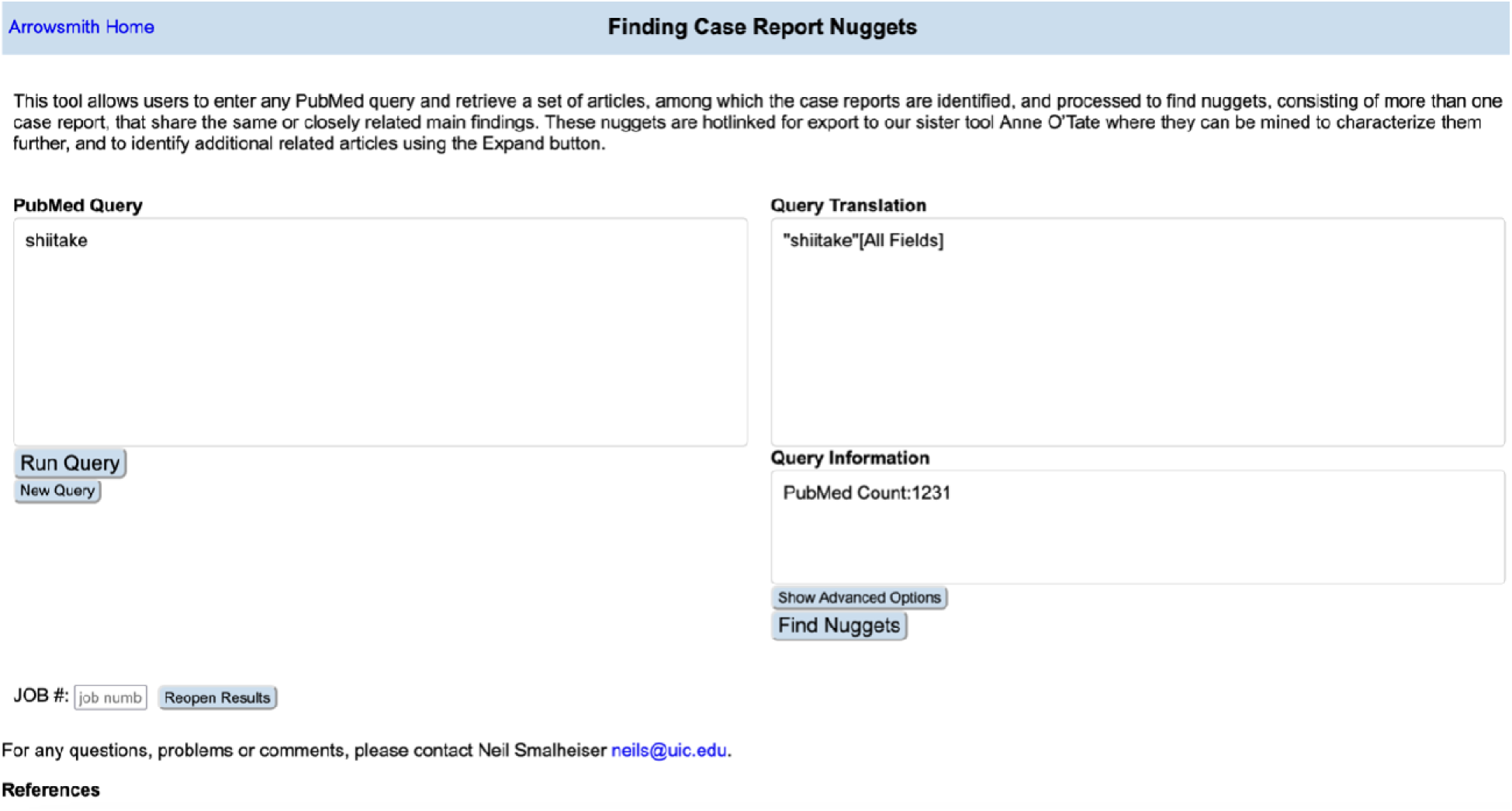
Home page of the Nuggets tool with user entering a search on “shiitake”.

**Figure 5.**
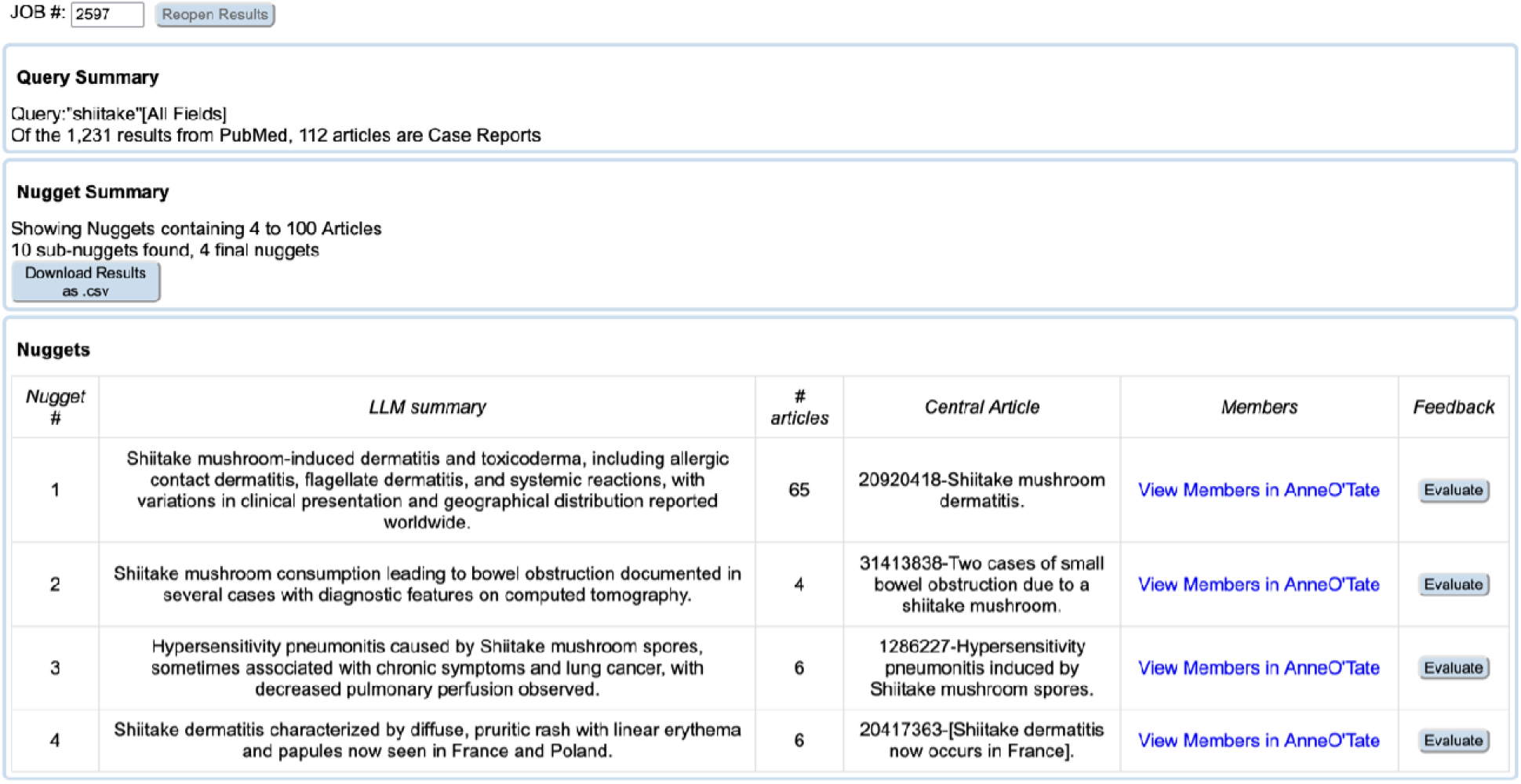
Output of the shiitake query identifies four nuggets.

**Figure 6.**
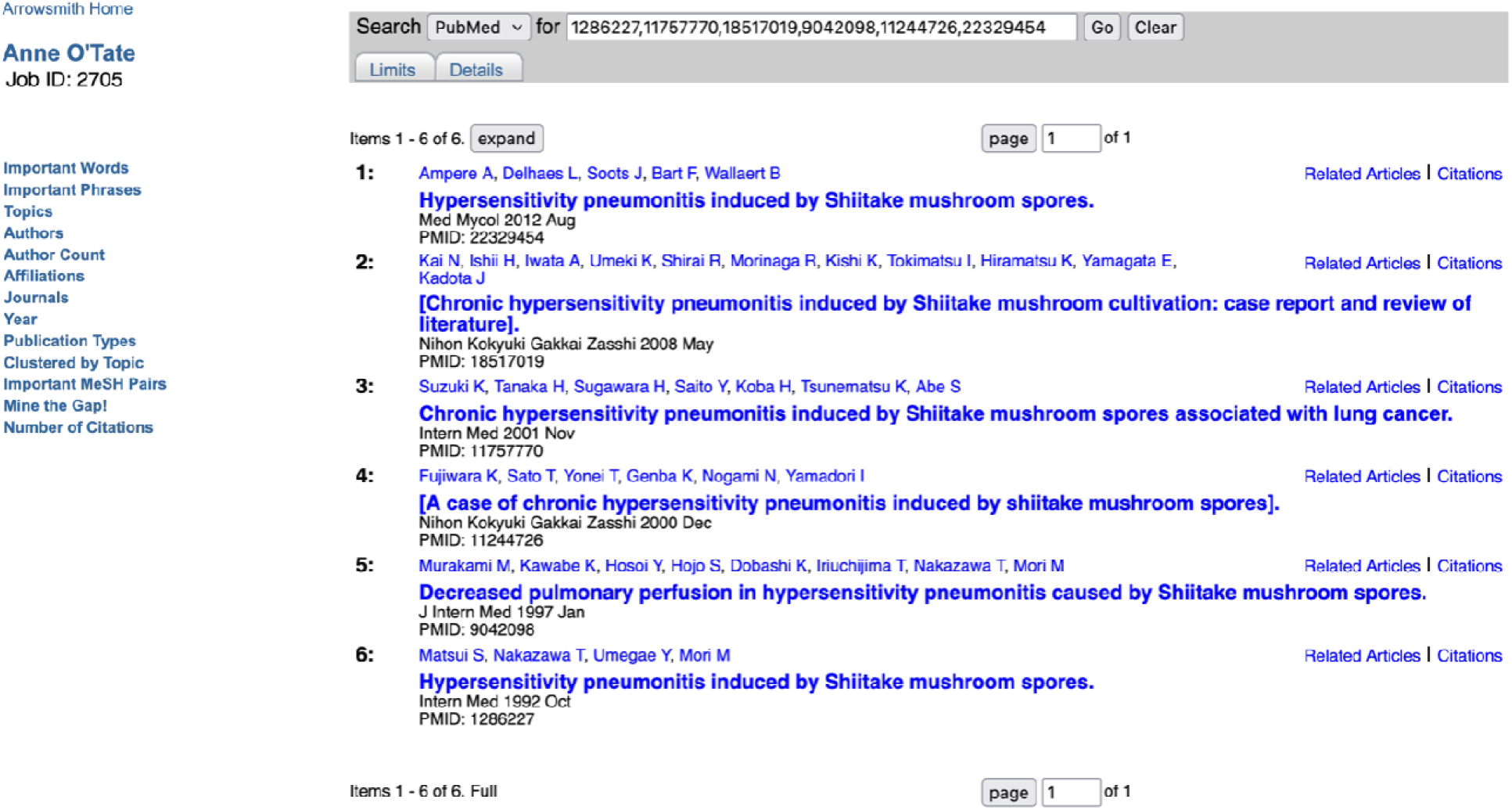
Nugget concerning shiitake hypersensitivity pneumonitis.

**Figure 7.**
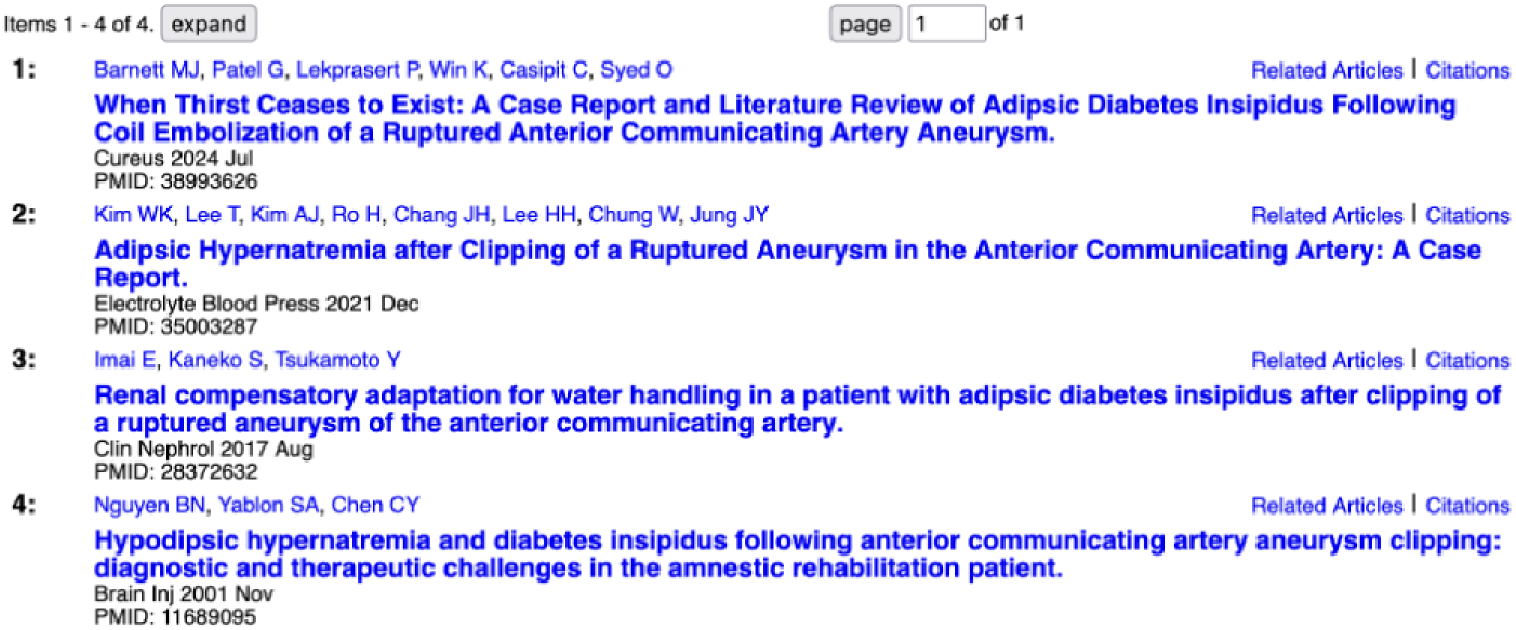
A PubMed query on lack of thirst identifies a single nugget containing four articles.

A second typical use case for the tool is to look for reports related to specific drugs, including but not limited to adverse drug effects. For example, for the popular drug zolpidem (brand name Ambien), the PubMed query “zolpidem“[MeSH Terms] OR “zolpidem“[All Fields] gives 2,980 results from PubMed, of which 410 articles are case reports. Our tool identifies 9 nuggets, of which 96 articles report on aspects of abuse and dependence of zolpidem, 21 report neuropsychiatric symptoms including psychosis, and others report miscellaneous but still well-documented phenomena, such as use of zolpidem to treat progressive supernuclear palsy and forms of dystonia.

A third example comes from a diagnostic dilemma. Suppose one has a patient who exhibits lack of thirst: What light can the case reports literature shine on possible causes? For the PubMed query [lack of thirst], there are 209 results, of which 31 articles are case reports, and only a single nugget consisting of 4 articles (Figure 7). All four describe diabetes insipidus and hypernatremia which followed clipping of a ruptured aneurysm in the anterior communicating artery – a rare and highly specific scenario, but one which is well documented in independent reports ranging from 2001 to 2024. Such a finding becomes important to consider in this patient population.

The tool offers several Advanced Options and allows users to download the results of an analysis, as well as to export a nugget to our Anne O’Tate tool for further mining and analysis. We encourage users to evaluate and give feedback on the quality of the nuggets that are displayed, by providing an “Evaluate” button next to each nugget which opens up in a pop-up and asks users to rate the nugget quality on a simple 1-3 Likert scale.

### Comparison with BioTextQuest v2.0

BioTextQuest v2.0 displays articles retrieved from PubMed separated into topical clusters.[35] A variety of options are available in terms of terms extracted (default = specialized terms vs. all text terms), similarity metrics (default = cosine vs. Jaccard vs. Euclidean), clustering algorithms (default = K-means vs. top2vec), and number of clusters (default set to 2). To examine whether such a tool would suffice to identify nuggets, using either default or custom parameters, we compared a number of PubMed queries to the Finding Case Report Nuggets tool vs. the BioTextQuest v2.0 tool. The differences may be exemplified using the query discussed above regarding lack of thirst. Querying [lack of thirst AND case reports[publication type]] into the BioTextQuest v2.0 tool using default parameters results in one cluster of 17 articles (containing only two of the articles identified as comprising a nugget) and another consisting of a single article. Setting the tool to extract all text terms identified one cluster of 27 articles (including all four of the articles in the nugget) and a second cluster of one article. Setting the number of clusters to 8 resulted in one big cluster of 21 articles (including all four of the articles in the nugget) plus six clusters containing one or two articles each. Raising the number of clusters to 16 resulted in splitting the nugget across two clusters (one containing 3 articles, the other containing the fourth). Thus, topical clustering did not readily capture the nugget identified by our tool.

## DISCUSSION

Many investigators have created biomedical data mining tools.[41] These are useful for e.g., identifying prominent themes or topics that are discussed in a set of articles retrieved from the biomedical literature, finding the most prolific authors, informative keywords, related articles, and so on. More recent LLM-based tools carry out more sophisticated tasks such as multi-document summarization.[42] Here we have created a more specific, specialized tool designed to identify nuggets – multiple clinical case reports that state the same, or almost the same, main findings. This builds upon our prior analyses showing that nuggets are surprisingly common within the case report literature, and that evidence reported in a nugget tends to be scientifically reliable and medically valuable.[19] Of course, the delineation of a nugget does not, by itself, provide evidence that a finding is scientifically valid, but it serves as a valuable pre-filter for manual inspection.

The **Finding Case Report Nuggets** tool is intended to complement, and not to replace, other types of literature analyses, since it aims for high precision and not for high recall. It does not attempt to find all case reports on the same topic, but to quickly find a core set of very similar reports that reinforce each others’ findings. The threshold of four articles to comprise a nugget was chosen because fewer than 0.2% of PubMed queries of geographical affiliations, that are topically diverse, gave rise to merged communities containing four or more articles (see Methods). Setting a threshold of four articles is a high bar that minimizes “noise” (though we do not mean to disparage the potential value of findings reported in only two or three articles, or even single case reports).

The tool may support at least four types of use cases:

● A clinician may have a patient with some unusual presentation and wants to learn quickly whether the case report literature has described similar findings repeatedly that might be relevant to their patient.
● A team may be compiling a systematic review on a particular topic. These are generally focused on controlled trials and controlled observational studies, but they might include case reports and case series if the latter provide sufficient credible evidence on that topic to be incorporated into the review. Our tool offers one strategy to assess this quickly. If a relevant nugget is identified, then the team would be motivated to look for additional articles using more comprehensive strategies such as following citation trails or making iterated PubMed queries. The tool does help in this regard too, for it allows the user to export the nugget to the Anne O’Tate tool where he or she can use the Expand button to identify additional similar articles, and the Citation Cloud function to identify citation trails.[40, 43]
● Although the tool is currently configured for public users to carry out specific PubMed searches on a given topic, it is also possible to configure the tool in the future to carry out automated surveillance of the biomedical literature, looking for one or more newly published case reports that reinforce previous reports, comprising a total of 4 or more articles and thus completing a nugget. This would be particularly valuable in detecting adverse effects of new therapies or emerging diseases.
● Finally, case reports are unique among biomedical articles in that the main finding is generally stated explicitly in the title.[19–21] For that reason, the titles of the different articles within a given nugget are largely paraphrases; thus, a corpus of nuggets may be useful as benchmarks for natural language processing models of semantic similarity.[44]

### Limitations

The nuggets tool has been designed in light of analyses of the case report literature, and of the prevalence and characteristics of nuggets observed within sets of articles retrieved from focused PubMed topical queries. However, not all articles have informative titles nor include abstracts, so some main findings may be missed. As well, the parameters for identifying nuggets, for example, the threshold for merging communities, have been assigned on an empirical basis. For that reason, results may not always be optimal for all queries. Similarly, the choice of resolution for the Louvain algorithm was also chosen on an empirical basis by examining to what extent well-defined communities are embedded within a dense network structure. In future research, we may seek to learn whether thresholds can be dynamically assigned based on the size and features of the set of retrieved articles. We also employ a popular, publicly accessible LLM to perform summaries of the titles comprising each nugget;[39] we have not formally checked these summaries for accuracy, coherence, consistency and clinical relevance, although there is a feedback button associated with each nugget so that users can submit feedback on the quality of the nuggets and their summaries.

## CONCLUSION

The **Finding Case Report Nuggets** tool (accessible at https://arrowsmith.psych.uic.edu/casereports) provides a web-based method for placing together multiple case reports with similar findings, offering a thumbnail sketch of a diverse literature and identifying nuggets that are especially likely to be credible. We hope that users will make use of the feedback button next to the displayed nuggets to help us to refine and improve the tool.

## Data Availability

An example query with underlying code has been deposited into DRYAD, DOI: 10.5061/dryad.5tb2rbpfb.

## ACKNOWLEDGMENTS

We thank Ang Michael Troy for assistance in evaluating nugget quality during the development of the tool.

## AUTHOR CONTRIBUTIONS

Neil Smalheiser: conceptualization, methodology, writing- original draft, writing -review and editing, supervision, funding acquisition.

Arthur Holt: formal analysis, investigation, writing - original draft, writing -review and editing, visualization.

## ETHICS APPROVAL AND CONSENT TO PARTICIPATE

Not applicable.

## CONSENT FOR PUBLICATION

Not applicable.

## AVAILABILITY OF DATA AND MATERIALS

An example query with underlying code has been deposited into DRYAD, DOI: 10.5061/dryad.5tb2rbpfb.

